# Detecting therapy-guiding RNA aberrations in platelets of non-small cell lung cancer patients

**DOI:** 10.1101/2021.01.26.21250013

**Authors:** Pei Meng, Anna A. Rybczynska, Jiacong Wei, Miente Martijn Terpstra, Wim Timens, Ed Schuuring, T. Jeroen N. Hiltermann, Harry J.M. Groen, Anthonie J. van der Wekken, Anke van den Berg, Klaas Kok

## Abstract

**Background:** Nowadays, the detection of therapy-guiding aberrations such as *EGFR* mutations and *ALK* fusion genes in tumor tissue is common practice for lung cancer patients. The aim of this study is to explore the feasibility of detecting common therapy-guiding aberrations in RNA isolated from platelets.

**Methods:** We applied a single primed enrichment-based targeted next generation sequencing (NGS) approach on 10 platelet RNA samples isolated from patients with active disease. In parallel, we applied RNA-based ddPCR focusing on *EGFR* p.(T790M), p.(L858R) and E19del, on *KRAS* codon 12 and 13 and on *ALK-EML4/KIF5B* fusions on 22 platelet RNA samples from patients with tumors known to harbor one of these drivers. For 11 cases ddPCR analysis of circulating cell free (cf)DNA from the same blood sample was performed in parallel.

**Results:** Despite having good quality NGS data, none of the variants detected in the tumor biopsy were observed in platelet-derived RNA samples. Using the more sensitive ddPCR, we detected known aberrations in three of 22 platelet-derived RNA samples. *EGFR* mutant droplets were not observed in any of the seven platelet samples, while these mutations were detected in three of six cfDNA samples of which five were extracted from the same blood sample. *KRAS* mutant droplets were detected in three out of nine platelet RNA samples with fractional abundances of 0.07%, 0.11% and 0.55%. Analysis of five cfDNA samples from the same blood samples revealed *KRAS*-mutant droplets in four of them. Two of the four corresponding platelet RNA samples were also positive for mutant *KRAS*. No *ALK* fusion droplets were detected in seven platelet derived RNA samples.

**Conclusions:** The level of tumor-derived RNA transcripts in platelets of non-small cell lung cancer patients was too low to be measured reliably for clinically relevant alterations in *EGFR, KRAS* and *ALK* fusion gene transcripts.

## Introduction

Most lung cancer patients present with advanced disease at diagnosis. Treatment decisions of advanced lung cancer patients have become largely dependent on genomic analyses, especially for adenocarcinoma [1,2]. Driver aberrations in *EGFR, KRAS* and *ALK* are observed commonly in adenocarcinoma of the lung. About 80% of the *EGFR* activating mutations are in frame deletions in exon 19, targeting codons 746 to 753, or a point mutation targeting codon 858 in exon 21. Several tyrosine kinase inhibitors (TKIs), such as erlotinib, gefitinib, afatinib, osimertinib, have been approved for treatment of patients carrying activating *EGFR* mutations [3,4]. ALK inhibitors such as crizotinib, alectinib, ceritinib, lorlatinib and brigatinib are approved for patients with *ALK* fusions either or not in combination with specific *ALK* resistant mutations. *KRAS* mutations in codon 12, 13 and 61 occur in 30% of all lung adenocarcinoma patients [5]. The presence of *KRAS* mutations is associated with resistance to EGFR, ALK and BRAF kinase inhibitors [6-10]. In general, *EGFR, ALK* and *KRAS* variations are mutually exclusive [11-14], although large cohort studies revealed a few tumors harboring concomitant variants [15,16].

Identification of therapy-guiding mutations and fusions have become routine practice for lung adenocarcinoma [17]. Sequencing tests are usually performed on DNA or RNA extracted from tumor biopsies. However, taking a tissue biopsy can cause substantial discomfort and be potentially risky for the patient. Moreover, re-biopsy of resistant or progressive lesions is often not possible due to the location and the frequently poor performance of patients with progressive disease. These hurdles may be overcome by using blood-based minimal invasive liquid biopsies including circulating tumor DNA (ctDNA), exosomes, platelet-derived RNA and circulating tumor cells (CTCs) [18]. A highly sensitive blood-based assay would be an ideal tool to monitor response to treatment and/or progression of disease.

The clinical application of exosomes and CTCs in non-small cell lung cancer (NSCLC) is still in an early stage [19,20]. ctDNA is released from tumor cells to the blood and thus potentially carries the genetic alterations of all tumor subclones [21,22]. The American Food and Drug Administration (FDA) has approved liquid biopsy companion diagnostics that uses a quantitative real-time polymerase chain reaction (qPCR) [23] or next-generation sequencing (NGS) test to identify *EGFR* mutations in cfDNA of metastatic NSCLC patients (https://www.fda.gov/medical-devices/vitro-diagnostics/list-cleared-or-approved-companion-diagnostic-devices-vitro-and-imaging-tools). In addition, other ctDNA-based assays are used to detect relevant predictive markers for NSCLC patients like Ageno-MassArray UltraSEEK [24] and Avenio Expanded ctDNA NGS [25]. Although there are some NGS assays, such as Avenio Expanded ctDNA NGS assay, that are able to detect *ALK* rearrangements, none of them is approved for clinical use.

Another source of tumor-derived molecules are blood platelets [26]. Platelets have been demonstrated to take up tumor-derived cytokines, transcripts whereas exosomes contain mRNA, microRNA and circular RNA [27]. Platelets in cancer patients are often referred to as tumor-educated platelets (TEPs) [28]. In a recent study, *EML4-ALK* gene fusions were successfully detected in platelets using reverse-transcription PCR (RT-PCR). This demonstrates the potential clinical value of analyzing platelet RNA for the presence of *ALK* fusion transcripts and thereby allows monitoring response to ALK inhibitors [29].

A potential advantage of platelet RNA as a source for tumor specific molecules is the higher amount of total RNA in comparison to the total amount of cell free (cf)DNA. In addition, tumor-derived RNA, albeit less stable, may be present in more copies than tumor-derived DNA. Together this suggests that platelets may be a better source to identify both tumor-derived gene fusions and mutations. If so, platelets would be an additional source for minimally invasive molecular testing for tumor specific abbreviations. We tested the most common therapy-driving variants in platelet RNA from NSCLC patients using our customized RNA-based NGS assay as described previously [30]. This assay showed high sensitivity to detect these variants even in FFPE tissue biopsies. In addition, we applied the more sensitive droplet digital (dd)PCR assay to detect *EGFR, KRAS* and *ALK* variations in platelet RNA. For a subset of the patients, we were able to also test cfDNA extracted from the same blood tube. To our knowledge, the feasibility of detecting *EGFR* and *KRAS* mutations and *ALK* fusions in platelet-derived RNA by both NGS and ddPCR is rarely explored.

## Materials and Methods

### Overview of samples blood samples

As part of the UMCG pathology biobank initiative, platelets and cell-free plasma from patients with lung cancer is routinely isolated from 4 ml blood as described previously [31], and stored at −80°C. For this study we obtained 25 platelet samples and platelet-free plasma samples from 20 patients with known driver mutations (Supplementary Table S1). The pre-treatment tumor biopsies of these patients carried 27 driver mutations including eight *EGFR*, nine *KRAS*, one *PIK3CA* mutation, six *ALK/EML4* fusions, two *ALK/KIF5B* fusions and one ROS1 fusion (Figure 1 and Supplementary Table S1). For 11 samples from eight patients cfDNA was available from the same blood tube. RNA isolated from platelets of 10 healthy subjects was used as control for threshold measurements of positive samples. RNA samples isolated from six cell lines carrying specific driver mutations, H1975 (*EGFR* p.(L858R), p.(T790M)), H1650 (*EGFR* E19del), H2228 (*ALK/EML4* E20/E6), A549 (*KRAS* p.(G12S)), KOPN8 (*KRAS* p.(G12D)) and HCT116 (*KRAS* p.(G13D)) were used as positive or negative controls for the indicated variants (see Supplementary Table S1). All patient samples were anonymized for the investigators. The study protocol complies to the Research Code of the University Medical Centre Groningen (https://www.rug.nl/umcg/research/documents/research-code-info-umcg-nl.pdf) and national ethical and professional guidelines (“Code of conduct; Dutch federation of biomedical scientific societies”, http://www.federa.org). The study ethics was approved by the OncoLifeS biobank committee (Biobank of cancer in UMCG). The biobank was approved by the UMCG research ethics committee. The study was conducted in accordance with the 1964 Helsinki declaration.

**Figure 1.**
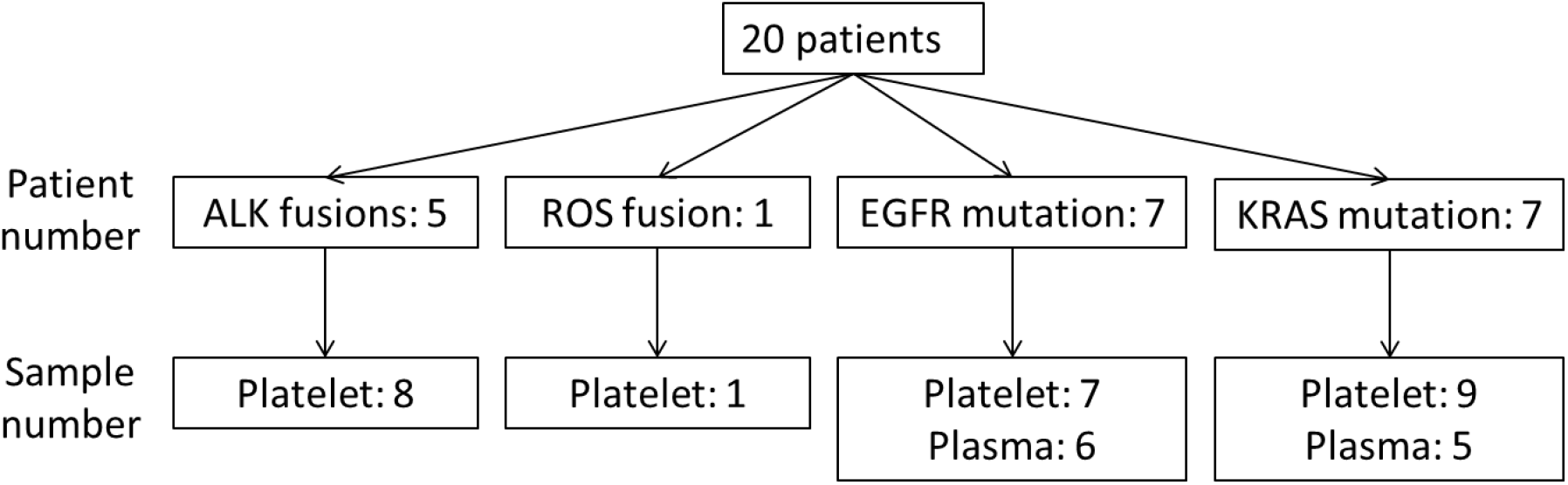
Overview of the selected samples. Indicated are the driver mutations detected in the tumor biopsy and the available platelet and plasma samples. One patient (P9) was diagnosed with both an *EGFR* E19del and an *EGFR* p.(T790M) mutation.

### RNA isolation and cDNA synthesis

Platelet RNA was isolated using the miRNeasy serum/plasma kit (Qiagen, Germany) and cDNA was synthesized using the IScript advanced cDNA synthesis kit (BIO-RAD Catalog# 1725037, USA). CfDNA was isolated from platelet-free plasma with the QIAamp Circulating Nucleic Acid Kit (Qiagen, Germany). RNA from cell lines was isolated using GeneJET RNA Purification Kit and cDNA was synthesized using the RevertAid H Minus First Strand cDNA Synthesis Kit (Thermo Fisher Scientific, USA). RNA concentrations were determined by NanoDrop 1000. The cDNA synthesis reaction for ddPCR was done using 145 ng to 2 μg RNA, dependent on the amount of available RNA, in a volume of 20 μl. All isolations and cDNA synthesis steps were done according to the manufacturers’ instructions.

### Targeted sequencing

The DV200 value, as determined by fragment analyzer (Agilent), was used as RNA-quality measurement. Libraries were prepared according to the user guide of the ovation fusion panel target enrichment system and ovation cDNA module for target enrichment (NuGEN) using the custom designed gene panel described elsewhere [30] with a total RNA input of 200-400 ng. Libraries were mixed in equimolar amounts and sequenced on the MiSeq platform (Illumina, San Diego, USA) with 150 bp paired-end reads according to the protocol of the manufacturer. As a quality marker for purity of the platelet samples, landing-probes for the platelet marker GP9 (platelet glycoprotein IX/CD42a) and the white blood cell marker PTPRC (CD45) were included in our targeted NGS design. Results obtained for the platelet samples were compared to our previous results for the same targeted NGS design on cell lines, pleural effusion, fresh-frozen and FFPE tissue samples [30]. In the current study we have applied the criteria for good quality data as determined in our previous study, i.e. DV200 >30 for the RNA sample, and >5⨯10^4^ unique sequence reads for a sample.

### DdPCR assay

Primers and probes for *EGFR* and *KRAS* mutation hotspots were ordered from Integrated DNA Technologies (IDT, Leuven, Belgium). Details about primers and probes for specific *EGFR* and *KRAS* variants are provided in Supplementary Table S2 [32]. The *KRAS* assays covered the seven most common *KRAS* G12/13 mutations. Probes specific for the mutations were labeled with Fluorescein amidites fluorescein (FAM), while all probes specific for wild-type sequences or for the control probe (in case for the *EGFR* E19del drop-off assay) were labeled with hexachlorofluorescein (HEX). The *ALK* fusion assay (Unique Assay ID dHsaEXD86850342, BIO-RAD, Hercules, CA, USA) covering the four most prevalent *ALK/EML4* fusion variants (GenBank Accession Number: AB274722.1, AB275889.1, AB374361.1, AB374362.1) all labeled with FAM were used in combination with GusB as house-keeping gene labeled with HEX. To enable the detection of the *ALK/KIF5B* fusion gene, an in house-designed primer for KIF5B was added to the *ALK* fusion assay (Supplementary Table S2). The modified assay was validated using RNA isolated from tumor tissue of the *ALK/KIF5B* fusion gene-positive patient.

The ddPCR reactions contained cDNA corresponding to 145 ng to 2 μg RNA for patient samples, 230 ng to 1.2 μg RNA for healthy individuals, and 0.05 ng to 190 ng RNA for cell lines, in 1× ddPCR Supermix for Probes (No dUTP), 900 nmol/L of each primer, and 450 nmol/L of each probe, which was divided over 3 to 7 wells. The reaction mix was prepared according to the manual of ddPCR Supermix for Probes (No dUTP). After adding 70 μL of droplet generation oil (Bio-Rad Laboratories), the cartridge was loaded to the Bio-Rad QX200™ Droplet Generator to generate droplets. After transfer of the droplets to a 96-well PCR plate, the plate was incubated at 95°C for 10 min, followed by 39 PCR cycles of 95°C for 30 sec, annealing for 60 sec and 72°C for 15 sec. Finally, the plate was incubated at 98°C for 10 min and cooled to 4 °C. Annealing temperatures were 59 °C for the *KRAS* and 55 °C for the *EGFR* and *ALK* assays. The temperature ramp change was 2 °C/sec for all steps. Droplets were counted on a Bio-Rad droplet reader (QX200) and analyzed with Bio-Rad QuantaSoft™ Analysis Pro. In each experiment, we included no-template controls (NTC) to identify environmental contamination, wild type (e.g. mutation-negative) controls and mutation positive controls. For *KRAS* p.(G12S/C/R), p.(G12D/A/V) and p.(G13D), we tested 9, 10, and 2 platelet RNA samples respectively from healthy individuals depending on the availability of RNA. For *EGFR* p.(L858R), E19del and p.(T790M), 2 platelet RNA samples from healthy individuals were tested. All controls and samples were analyzed in at least three independent wells.

For each assay, optimal experimental conditions were established by a gradient annealing temperature on a positive and a negative control sample. To determine the lower limit of detection, the starting amount of cDNA input of positive control samples was adapted to a starting fractional abundance of 8% for *KRAS*, 4% for *EGFR*, and a mutant droplet number of 128 for the *ALK* fusion. Next, a 2-fold dilution series of the positive control sample was made in duplicate and analyzed. The threshold to discriminate between positive and negative droplets was set in the middle of positive and negative droplets of control samples except for the *EGFR* E19del drop-off assay for which we used a lower threshold.

## Results

### Targeted NGS of platelet samples

The DV200 value of the platelet-derived RNA samples ranged from 49-76 and fulfilled the cut-off criterium of 30 as set for our assay [30]. The number of unique on-target reads ranged from 1.2⨯10^5^ to 1.4⨯10^6^ (mean 8.4⨯10^5^), again well above 5⨯10^4^ which is the threshold determined in our previous study (Figure 2). The number of unique on-target reads was comparable to that obtained for cell lines and pleural effusions in our previous study [33] (Figure 2), which indicated that the quality of the RNA samples was good.

**Figure 2.**
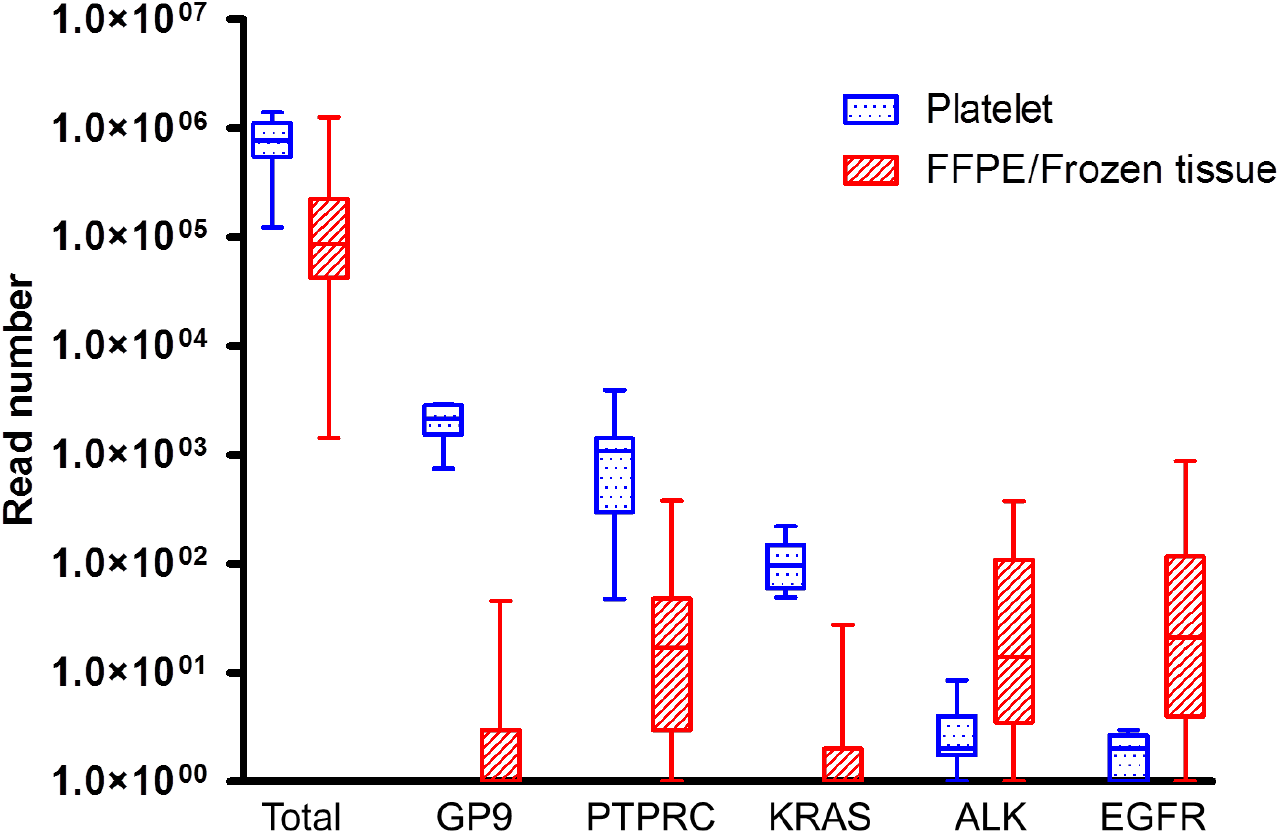
Unique on-target read counts obtained for 10 platelet samples and 53 fresh frozen or FFPE samples of cell lines, pleural effusions and biopsies. Platelets show a high read count for the platelet-specific transcript GP9 as compared to the non-platelet samples. Overall, GP9 reads were higher than the WBC marker PTPRC reads in our platelet samples. Platelets are characterized by a high abundance of wild type *KRAS* and a low abundance or even absence of *EGFR* transcripts.

The number of reads for the platelet marker GP9 was as expected highly abundant for the platelet samples and low for the cell lines, pleural effusions and tissue samples from our previous study (Figure 2). Moreover, GP9 read counts were also much higher than the read counts for the WBC marker PTPRC in our platelet samples, indicating efficient enrichment of platelet-derived RNA, with limited contamination of white blood cell-derived RNA. We observed high *KRAS* read counts in platelet RNA samples of patients and controls, indicating that platelets of both healthy controls and lung cancer patients contain high levels of *KRAS* mRNA. In contrast, overall *EGFR* read counts were very low, or even undetectable (Figure 2) in platelets from both patients and healthy controls. The somatic mutations for *EGFR* (n=3) and *KRAS* (n=4) identified in the tumor biopsies, were not detected in the corresponding platelet samples. Also, the ROS1 fusion, the *PIK3CA* mutation, and the two *ALK* fusions were not detected by our targeted NGS-based assay.

### Mutation and fusion gene detection by ddPCR

Based on the negative findings by our targeted NGS approach, we switched to the more sensitive ddPCR assay. After optimizing conditions for each ddPCR assay, a 2-fold dilution series was made from a positive control cell line. This revealed a lower limit of detection of four copies per well with a fractional abundance down to 0.03% (Supplementary Figure S1). For platelet RNA from healthy individuals containing a high number of wild-type *KRAS* transcripts, the average number of false positive mutant *KRAS* droplets was less than two per well (Supplementary Figure S2). Based on these observations, we considered patient samples to be true positive for *KRAS* when at least two positive droplets were observed in each single well and when the mutant droplet count of the negative-control cell line and the no-template control were less than two in the same run. The same criteria were adopted for the *EGFR* and *ALK* assays.

Analysis of seven platelet samples that were derived from four patients with a previously confirmed *ALK/EML4* or *ALK/KIF5B* fusion gene in their primary tumor, revealed no *ALK* fusion positive droplets (Figure 3). In the same analyses, the number of droplets for the GusB housekeeping gene ranged from 844 to 11,023, which was in the same range as observed in the *ALK* fusion positive and negative control samples.

**Figure 3.**
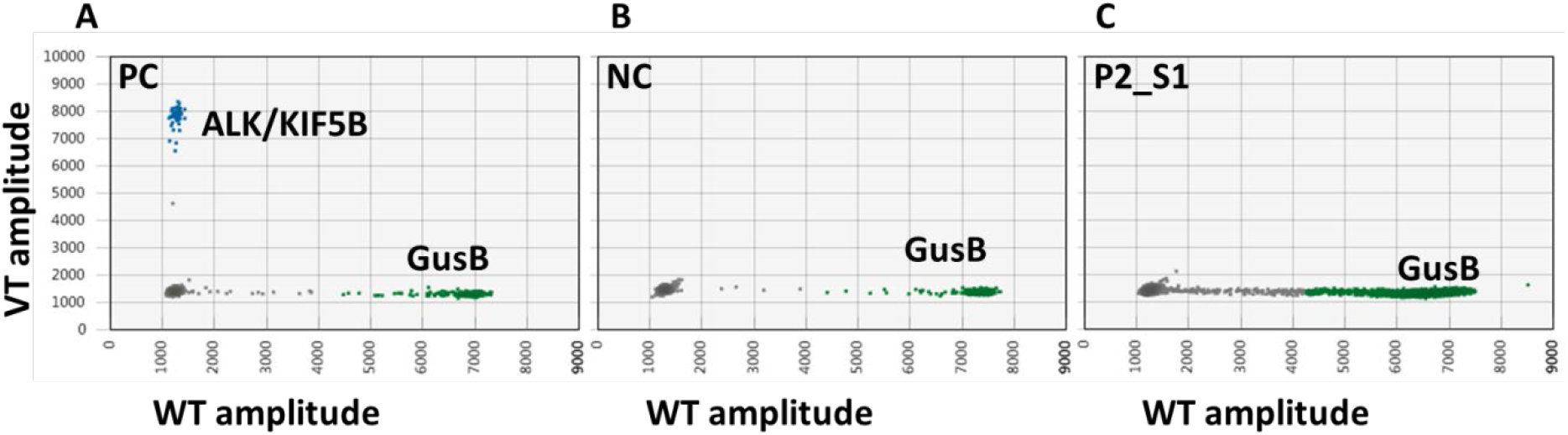
Representative ddPCR assay for the *ALK/KIF5B* fusion transcript. A: ddPCR of an FFPE tumor sample of a patient with an *ALK/KIF5B* fusion transcript. B: The A549 cell line was included as a negative control. C: Platelet sample of the patient with the *ALK/KIF5B* fusion transcript that was determined in the corresponding tissue sample (shown in panel A). No *ALK/KIF5B* fusion transcript-positive droplets (in blue) were detected, while a high number of house-keeping gene (GusB) droplets (in green) were detected. PC=positive control; NC=negative control

*EGFR* wild type droplets were virtually absent in platelet samples from six patients and two healthy individuals. This was consistent with the low read counts obtained in the targeted NGS assay and indicates a very low abundance of *EGFR* transcripts in both platelets from healthy controls and NSCLC patients. Consistent with the lack of wild type *EGFR* droplets, we also did not observe mutant *EGFR* droplets in platelet samples from the six patients with known *EGFR* variants, including three *EGFR* E19Del and three p.(L858R) variant samples (Figure 4).

**Figure 4.**
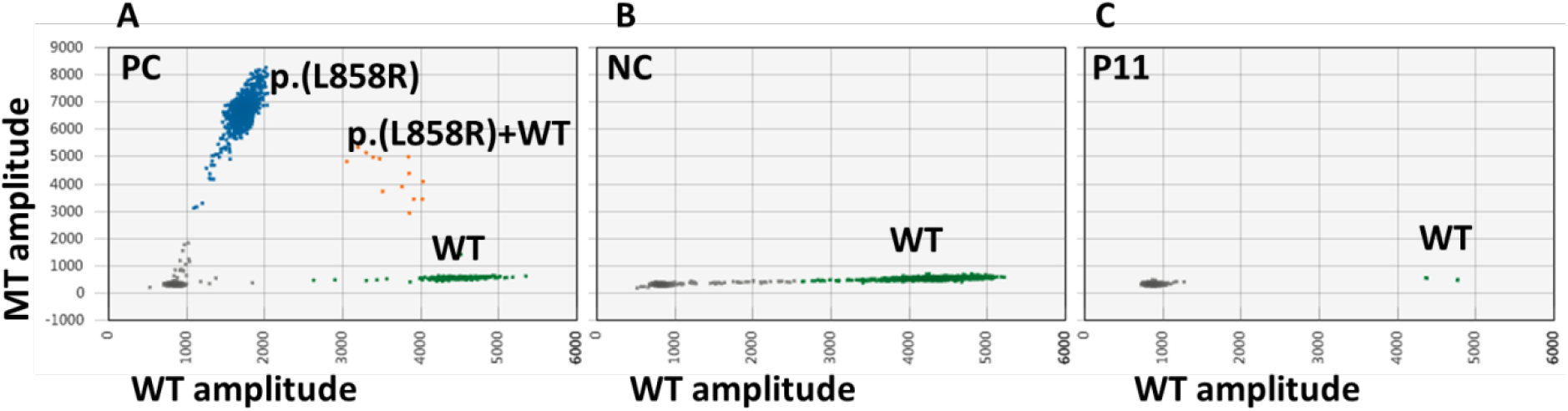
Representative ddPCR assay for the *EGFR* p.(L858R) mutation. A: The H1975 cell line with a p.(L858R) mutation was used as a positive control. B: The H1650 cell line was used as a negative control. C: Platelet sample from a patient with a p.(L858R) mutation in corresponding tumor tissue. No *EGFR* mutant droplet and only a few wild type *EGFR* droplets were detected in the platelet sample of this patient. The number of WT *EGFR* droplets was very low for all platelet samples derived from patients and healthy individuals, indicating a virtual absence of endogenous *EGFR* transcripts in platelets. WT: wild type *EGFR* droplets.

*KRAS* wild type droplet counts were very high in platelet samples from seven patients and 10 healthy individuals, with positive droplet counts ranging from 2,091 to 40,185 and from 7,877 to 26,926, respectively (numbers refer to the sum of all wells analyzed per sample). This was consistent with the NGS data, where a high number of wild type *KRAS* reads was detected in all platelet-derived RNA samples. *KRAS* p.(G12D) or p.(G13D) positive droplets were detected in 3 out of 9 platelet samples from patients carrying this mutation in their corresponding tumor, with fractional abundances of 0.07%, 0.11% and 0.55%. Two of the positive platelet samples were derived from one patient, obtained at progression to different treatments (Figure 5). The *KRAS* mutant droplet counts for all patient and healthy individuals are shown in supplementary figure S2.

**Figure 5.**
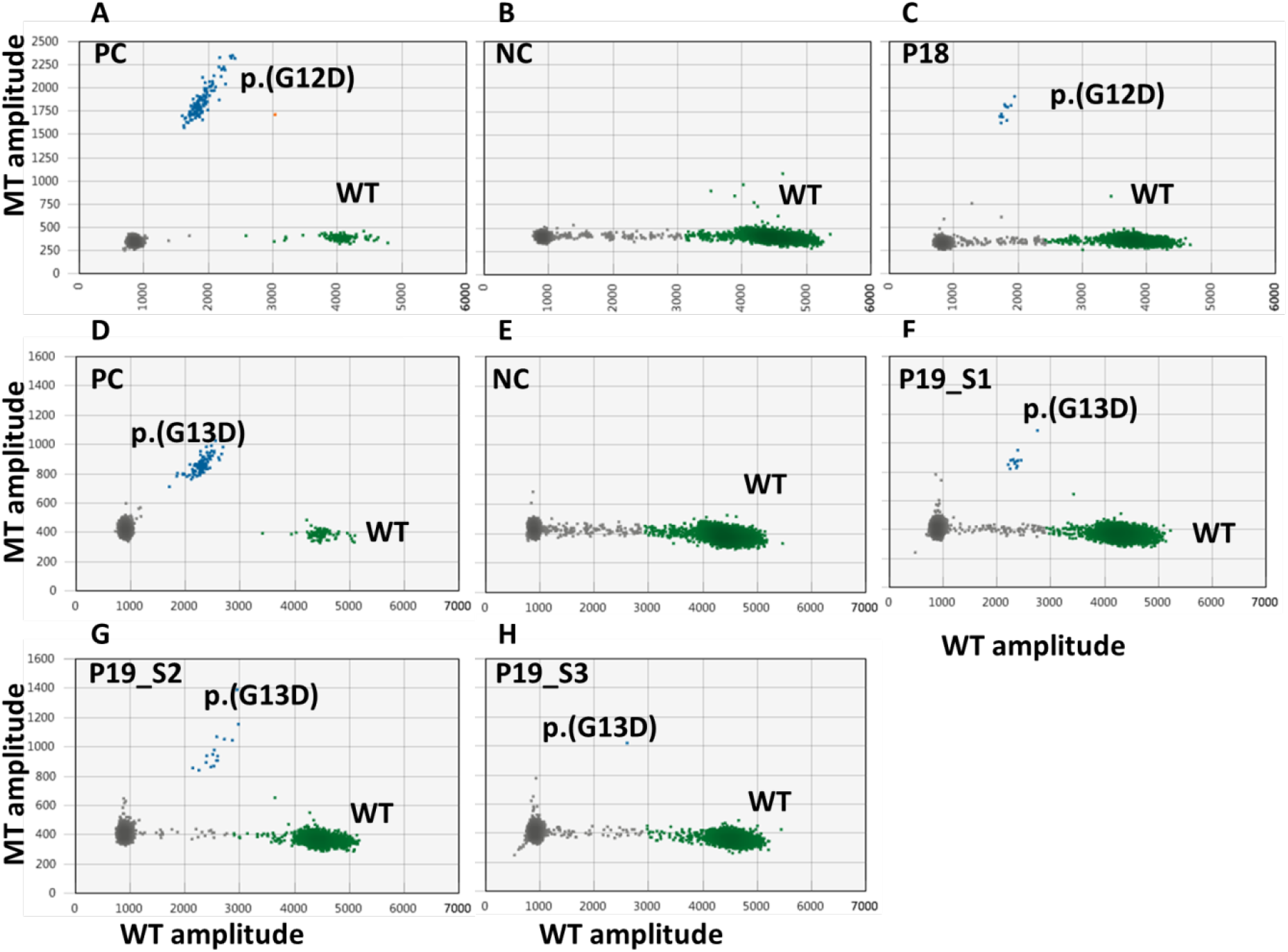
Representative ddPCR assays for *KRAS* p.(G12D) and p.(G13D) mutations. A: The KOPN8 cell line was used as a positive control for the *KRAS* p.(G12D) mutation. B: The H1975 cell line was used as a negative control. C: Platelet sample of a patient with a previously proven *KRAS* p.(G12D) mutation before treatment/at baseline. D: The HCT116 cell line was used as a positive control for the *KRAS* p.(G13D) mutation. E: The H1975 cell line was used as a negative control. F, G, H: Platelet samples of a single patient (P19) with a *KRAS* p.(G13D) mutation obtained at progression (F, G) or stable disease (H). The *KRAS* mutation was detected at a low fractional abundance in two of the three samples. WT: wild type *EGFR* droplets.

### Fractional abundance of *EGFR* and *KRAS* mutations in platelet RNA and cfDNA

For six *EGFR* and five *KRAS* mutation-positive patients (one patient had both an *EGFR* E19del and a p.(T790M) mutation), we had both platelet derived RNA and plasma-derived cfDNA from the same blood tube. Mutant droplets for the E19del were observed at a frequency of 3% and 37% in two of the three cfDNA samples. For the p.(L858R) positive patient the variant droplet frequency was 23% in the cfDNA, and for the p.(T790M) positive patient the variant droplet frequency was 18.4%. No mutant droplets were detected in the corresponding platelet RNA samples. In four of the five *KRAS* mutant-positive cfDNA samples, fractional abundances ranging from 8% to 20% were observed. In corresponding platelet samples of two of these patients we also observed the *KRAS* mutation, albeit at lower frequencies, i.e. 0.11% versus 8% and 0.55% versus 20% (see details in Table 1). For the fifth patient, with stable disease during treatment, both the cfDNA and the platelet RNA sample were negative.

**Table 1.** Overview of the ddPCR results of paired platelet derived RNA and plasma derived cfDNA samples.

| Gene | AA change | Fractional abundance (%) |  |  |
| --- | --- | --- | --- | --- |
|  |  | Targeted NGS | ddPCR |  |
|  |  | platelet RNA | platelet RNA | cfDNA |
| *EGFR* | E19del* | NA | 0 | 3 |
|  |  | NA | 0 | 0 |
|  |  | 0 | NA | 37 |
|  | L858R | NA | 0 | 0 |
|  |  | 0 | 0 | 23 |
|  |  | 0 | 0 | 0 |
|  | T790M | 0 | NA | 18.4 |
| *KRAS* | G12A | 0 | 0 | 0 |
|  | G12C | 0 | 0 | 10 |
|  | G13D | 0 | 0.11 | 8 |
|  |  | NA | 0.55 | 20 |
|  |  | NA | 0 | 14 |

\* One sample had both *EGFR* E19del and T790M. NA; not applicable.

## Discussion

Tumor cells can release RNA into the circulation via a variety of microvesicles [34,35], which in turn can be absorbed by platelets. These so-called tumor-educated platelets (TEPs) are thought to offer an alternative, less invasive source of tumor-derived RNA for the detection of oncogenic mutations. In the present study, we investigated the feasibility of using platelet-derived RNA to detect both SNVs and fusion genes relevant for treatment in advanced lung adenocarcinoma patients. Using our recently published RNA-based targeted NGS approach, we did not detect any of the expected 12 driver mutations in platelet RNA of patients with active disease, despite having good quality RNA (DV200 >30) and a high number of unique reads. The high read counts for the platelet-specific marker GP9 and the much lower read counts for the leukocyte specific marker PTPRC (CD45) indicated that the RNA indeed originated from platelets [36]. Subsequent follow-up by ddPCR revealed presence of mutant droplets only for three out of nine *KRAS*, and no mutant positive droplets for *EGFR* and *ALK* fusion gene transcripts.

A large RNA-seq study on platelet RNA derived from 228 patients with different tumor types and 55 healthy individuals revealed that using targeted ultra-deep sequencing (∼5,000× coverage), mutations were detected in 7/26 (26.9%) *KRAS* mutated patients and 4/11 (36.4%) *EGFR* mutated patients [28]. However, analysis of platelets derived from wild type samples revealed false positive results for *KRAS* and *EGFR* in 5/41 (12.2%) and 3/37 (8.1%) cases, respectively. In another study, *ALK-EML4* fusion transcripts were detected in platelets of 22 of 34 (64.7%) *ALK-EML4* positive NSCLC patients by RT-PCR. However, the authors used 60 PCR cycles to demonstrate the fusion genes. Thus, it appears that the fusion transcripts were present at extremely low levels [37]. Together with our data this suggests that the abundance of tumor cell-derived transcripts for *KRAS, EGFR* and *ALK* fusion genes are extremely low in platelet derived RNA sample. This does not rule out the potential value of platelet derived RNA as disease biomarkers. In recent years, TEPs have been recognized as a source of biomarkers to distinguish healthy individuals from individuals with active disease. The RNA splicing products present in TEPs have been reported to have diagnostic potential as pan-cancer, multiclass cancer, and companion diagnostics [28,38]. Decreased *ITGA2B* mRNA levels have been reported in platelets of patients with stage I NSCLC and this was suggested to be a promising biomarker to differentiate malignant from benign lung nodules [39]. More recently, expression levels of an 11-gene panel that included *ITGA2B* in platelets was shown to identify NSCLC patients with a high accuracy [40]. These latter studies clearly show altered RNA content in platelets of patients with cancer.

The high number of wild type transcripts in platelet samples preclude reliable detection of tumor derived mutant transcripts, which are present at very low copy numbers. A comparative study detected *BRAF* p.(V600E) in 10 of 12 RNA samples from extracellular vesicles in melanoma patients [41]. However, none of the *BRAF* mutations were detected in platelet-derived RNA samples of these 12 patients while high amounts of wild-type BRAF transcripts were detected. This study concluded that *BRAF* p.(V600E) mutant transcripts are too rare in the platelet RNA fraction to be reliably detected [41]. So, these findings are similar to our findings for KRAS and both suggest low abundance of tumor derived transcripts in platelets. In contrast to *KRAS* and *BRAF*, the amount of wild type *EGFR* transcripts was very low in all platelet samples, irrespective of being derived from patients or healthy individuals. No *EGFR*-mutant droplets were observed in platelets obtained from *EGFR* mutation positive patients. These data also indicate lack of tumor cell derived *EGFR* transcripts in platelets, consistent with *KRAS* and *BRAF*.

It has been shown that NSCLC driver mutations can be detected in cfDNA of patients with active disease. This enables testing *EGFR* mutations that guide EGFR inhibitor therapy without the need of a tissue biopsy in advanced NSCLC patients [42]. Indeed, in our study, ddPCR analysis of cfDNA isolated from the same blood samples from which we derived the platelets revealed the presence of the expected driver mutations in seven out of eleven cases. For four of these seven cases we failed to detect mutant-positive droplets for our platelet-derived RNA-based assay. A disadvantage of the application of cfDNA is that the screening for *ALK* fusions is more challenging, although not impossible [43]

Taken together, our results indicate that the proportion of tumor derived RNA molecules in platelets is extremely low. *EGFR* and *KRAS* mutation-positive droplet frequencies detected in cfDNA are much higher than detected in platelets and thus represents a more reliable source to study disease dynamics.

## Conclusions

Our study showed that the amount of tumor-derived RNA transcripts in platelets of NSCLC patients are too low to determine alterations in *EGFR, KRAS* and *ALK* transcripts with a sensitivity that is needed in a diagnostic setting.

## Data Availability

I have followed all appropriate research reporting guidelines and uploaded the relevant EQUATOR Network research reporting checklist(s) and other pertinent material as supplementary files, if applicable.

## Funding

This research was funded by the Dutch Cancer Society Grant: KWF grant (RUG2015-8044).

## Acknowledgments

We would like to acknowledge the molecular diagnostics team of the Department Pathology and the sequencing team of the Department of Genetics for technical assistance with the experimental work. We would like to thank the Centre for Information Technology of the University of Groningen for their support and for providing access to the Peregrine high-performance computing cluster.

## Conflicts of Interest

The authors declare no conflict of interest.

Ed Schuuring has performed lectures for Bio-Rad, Novartis, Roche, Biocartis, Illumina, Pfizer, AstraZeneca, and Agena Bioscience, is consultant in advisory boards for AstraZeneca, Roche, Pfizer, Novartis, Bayer, Lilly, BMS, Amgen, BioCartis, Illumina, Agena Bioscience and MSD/Merck, and received research grants from Pfizer, Biocartis, Agena Bioscience, BMS, Bio-Rad, Roche, Boehringer Ingelheim. Wim Timens reports fees to Institution for lectures or consultancy from Roche Diagnostics / Ventana, Merck Sharp Dohme, Bristol-Myers-Squibb, and AbbVie, outside the submitted work. Harry J M Groen reports grants from Boehringer-Ingelheim, Takeda, BMS, Novartis, Merck outside the submitted work. Anthonie van der Wekken has performed lectures for Lilly, Boehringer-Ingelheim, Pfizer, AstraZeneca, Roche (diagnostics) and Takeda, is consultant in advisory boards for Lilly, Boehringer-Ingelheim, Pfizer, AstraZeneca, Roche (diagnostics), Takeda and Janssen, received grants from Boehringer-Ingelheim, Pfizer, AstraZeneca, Roche and Takeda all outside the submitted work. Jeroen Hiltermann reports grants from AstraZeneca, Roche, BMS, outside the submitted work.

## Notes

### Author Declarations

The study ethics was approved by the OncoLifeS biobank committee (Biobank of cancer in UMCG). The biobank was approved by the UMCG research ethics committee. The study was conducted in accordance with the 1964 Helsinki declaration.

