## Supplementary material for "Detecting therapy-guiding RNA aberrations in platelets of non-small cell lung cancer patients": supplementary.docx

The Netherlands

**Supplementary data**

**Supplementary Table S1.** Testing of platelet RNA and plasma cfDNA variants overview

| Patient ID | MD aberration | | | Targeted RNAseq_ platelet RNA | | ddPCR_platelet RNA | | ddPCR_cell free DNA | | Time point of the sample |
| --- | --- | --- | --- | --- | --- | --- | --- | --- | --- | --- |
|  | Gene | CDS/breakpoint | AA change | ALT | VAF (%) | variant droplets* | Fractional abundance (%) | variant droplets | Fractional abundance (%) |  |
| P1 | *ALK* | *ALK/EML4* | NA | 0 | 0 |  |  |  |  | at progression |
| P2_S1 |  | *ALK/KIF5B* | NA | 0 | 0 | 0 | 0 |  |  | at progression |
| P2_S2 |  | *ALK/KIF5B* | NA |  |  | 0 | 0 |  |  | at progression |
| P3 |  | *ALK/EML4* | NA |  |  | 0 | 0 |  |  | at progression |
| P4_S1 |  | *ALK/EML4* | NA |  |  | 0 | 0 |  |  | at progression |
| P4_S2 |  | *ALK/EML4* | NA |  |  | 0 | 0 |  |  | at progression |
| P5_S1 |  | *ALK/EML4* | NA |  |  | 0 | 0 |  |  | at progression |
| P5_S2 |  | *ALK/EML4* | NA |  |  | 0 | 0 |  |  | at progression |
| P6 | *EGFR* | 2235_2249del | E746_A750del |  |  | 0 | 0 |  |  | at diagnosis |
| P7 |  | 2235_2249del | E746_A750del |  |  | 0 | 0 | 17 | 3 | at progression |
| P8 |  | 2240_2257del | L747_P753delinsS |  |  | 0 | 0 | 0 | 0 | at progression |
| P9 |  | c.2236_2250del | E746_A750del | 0 | 0 |  |  | 184 | 37 | at progression |
| P10 |  | 2573 T>G | L858R |  |  | 0 | 0 | 0 | 0 | at progression |
| P11 |  | 2573T>G | L858R | 0 | 0 | 0 | 0 | 296 | 23 | at diagnosis |
| P12 |  | 2573T>G | L858R | 0 | 0 | 0 | 0 | 0 | 0 | at diagnosis |
| P9 |  | 2369C>T | T790M | 0 | 0 |  |  | 14 | 18.4 | at progression |
| P13 | *KRAS* | 35G>C | G12A | 0 | 0 | 0 | 0 | 1 | 0 | at stable disease |
| P14 |  | 34G>T | G12C | 0 | 0 | 0 | 0 | 225 | 10 | at progression |
| P15 |  | 34G>T | G12C | 0 | 0 | 0 | 0 |  |  | at progression |
| P16 |  | 34G>T | G12C |  |  | 0 | 0 |  |  | at progression |
| P17 |  | 34G>T | G12C |  |  | 0 | 0 |  |  | at progression |
| P18 |  | 35G>A | G12D |  |  | 10 | 0.07 |  |  | at diagnosis |
| P19_S1 |  | 38G>A | G13D | 0 | 0 | 13 | 0.11 | 18 | 8 | at progression |
| P19_S2 |  | 38G>A | G13D |  |  | 16 | 0.55 | 1172 | 20 | at progression |
| P19_S3 |  | 38G>A | G13D |  |  | 0 | 0 | 509 | 14 | at stable disease |
| P12 | *PIK3CA* | 1673A>G | Q546R | 0 | 0 |  |  |  |  | at diagnosis |
| P20 | *ROS1* | unknown | NA | 0 | 0 |  |  |  |  | at progression |

*The variant droplet number are present as 0 when the average droplet number is less than 2 in each well.

**Supplementary Table S2.** Overview of the Primers and Probes for the ddPCR Assays

| **Gene** | **Mutation** | **Primer/Probe** | **Sequence 5'-3'** | **Dye** | **Length** | **GC%** | **Tm(℃)** | **Amplicon** |
| --- | --- | --- | --- | --- | --- | --- | --- | --- |
| *KRAS* | G12/G13 | FP | TAAGGTGCGGGAGAGAGG |  | 18 | 61 | 56 | 97 bp |
|  |  | RP | TAGCTGTATCGTCAAGGCAC |  | 20 | 50 | 54 |  |
|  |  | WT probe | TTGGAGCTGGTGGCGT | HEX | 16 | 63 | 57 |  |
|  | G12S/R/C | Mut probe | TTGGAGCT[A/C/T]GTGGCGT | FAM | 16 | 58 | 55 |  |
|  | G12D/A/V | Mut probe | TGGAGCTG[A/C/T]TGGCGT | FAM | 15 | 62 | 55 |  |
|  | G13D | Mut probe | CTGGTGACGTAGGCA | FAM | 15 | 60 | 51 |  |
| *EGFR* | T790M | FP | CAAGGAAATCCTCGATGAAGCC |  | 22 | 50 | 55.9 | 154 bp |
|  |  | RP | GTCTTTGTGTTCCCGGACATAGT |  | 23 | 48 | 56.9 |  |
|  |  | WT probe | ATGAGCTGCGTGATGAG | HEX | 17 | 53 | 52.2 |  |
|  |  | Mut probe | ATGAGCTGCATGATGAG | FAM | 17 | 47 | 49.2 |  |
|  | L858R | FP | GCAGCATGTCAAGATCACAGATT |  | 23 | 44 | 55.7 | 100 bp |
|  |  | RP | CATCCACTTGATAGGCACTTTGC |  | 23 | 48 | 56.4 |  |
|  |  | WT probe | AGTTTGGCCAGCCCAA | HEX | 16 | 56 | 54.9 |  |
|  |  | Mut probe | AGTTTGGCCCGCCCAA | FAM | 16 | 63 | 58.2 |  |
|  | delE19 | FP | GTGAGAAAGTTAAAATTCCCGTC |  | 23 | 39 | 52.1 | 97 (82) bp |
|  |  | RP | TGGCCATCACGTAGGCTTC |  | 19 | 58 | 57.5 | 97 (82) bp |
|  |  | Common probe | ATCGAGGATTTCCTTGTTGGCT | HEX | 22 | 46 | 56.6 |  |
|  |  | Mut probe | AAGGAATTAAGAGAAGCAACATCTCC | FAM | 26 | 39 | 55.1 |  |
| *KIF5B*_e27_FP* | *ALK/KIF5B* | FP | AGGGCATTCTGCACAGATTG |  | 20 | 50 | 55.7 | > 90 bp |

FP: forward primer, RP: reverse primer, WT: wild type, Mut: mutant type; *This FP was directly added to the commercial *ALK* assay with probes and primers.

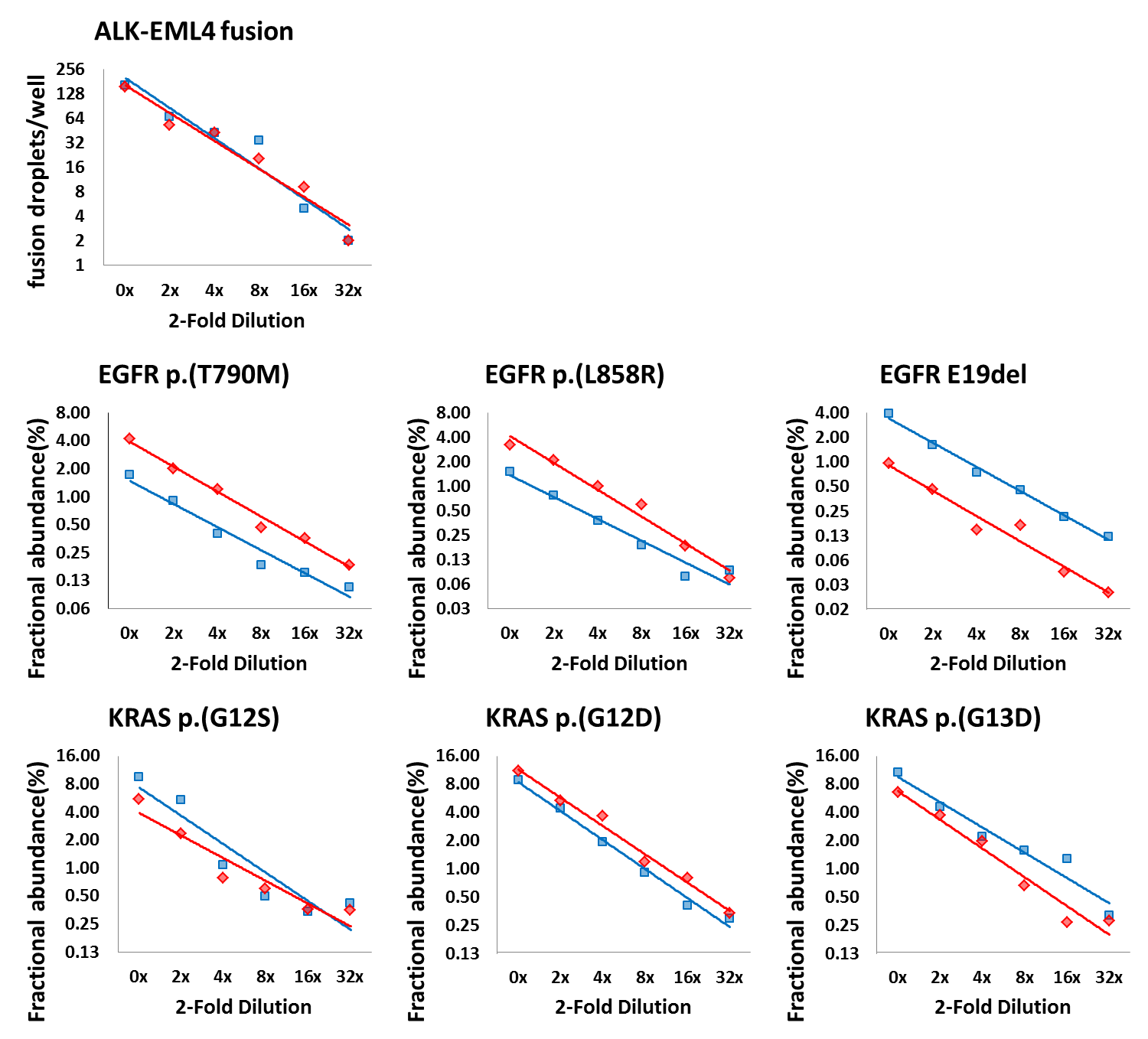

**Supplementary Figure S1.** *ALK-EML4* fusion gene, and *EGFR* and *KRAS* mutation detection by RNA-based ddPCR in a 2-fold dilution series of a mutation-positive control cell line. Each line represents one of the two independent 2-fold dilution series. The number of *ALK-EML4* fusion droplets ranged from 2 to 163 and 2 to 153. The *KRAS* fractional abundances ranged from 0.42% to 9.53% and 0.35% to 5.54% (G12S), 0.34% to 11.16% and 0.30% to 8.97% (G12D), 0.28% to 6.56% and 0.33% to 10.80% (G13D). The *EGFR* fractional abundances ranged from 0.11% to 1.79% and 0.18% to 4.14% (T790M), 0.09% to 1.5% and 0.07% to 3.22% (L858R), 0.03% to 0.95% and 0.12% to 3.86% (Exon 19 del).

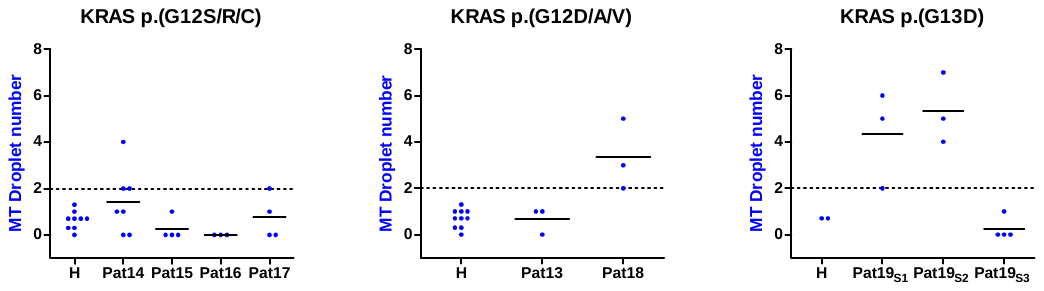

**Supplementary Figure S2.** Summary of *KRAS* mutant droplet numbers in platelet RNA from patients and healthy individuals. For the healthy individuals each dot indicates the average of a threefold measurement. For patient samples, each dot indicates a single measurement. H: healthy individuals.
